# Spatiotemporal Interactions of Air Pollution, Airborne Pollen, and Land Cover on Asthma and Allergies Medication Sales: A Population-Level Ecological Study

**DOI:** 10.64898/2026.07.20.26358483

**Authors:** Isabella Annesi-Maesano

**Author notes:** Corresponding author: IAM.

## Abstract

**Introduction:** The interaction between air pollution and airborne pollen is emerging as a major public health concern, as it may enhance allergic responses and exacerbate asthma at the population level. Available evidence suggests that urban residents experience a higher burden of respiratory allergies than rural populations, potentially due to the combined effects of chemical air pollutants and pollen exposure. Air pollutants may modify pollen characteristics, increase allergen release and potency, and promote airway inflammation, thereby amplifying allergic sensitization and respiratory symptoms.

**Aims:** For the first time, the relationship between air pollution, pollen, and asthma and allergies is investigated by simultaneously factoring in spatiotemporal land cover data (urban, agricultural, and forest spaces).

**Methods:** We utilized descriptive statistics, Principal Component Analysis (PCA), Hierarchical Cluster Analysis (HCA), and spatial analysis to understand the relationship of birch, grass and all taxons count, NO_2_ and PM_2.5_ concentrations (micro grams/m^3^) and land cover type (urban/rural, agriculture, forest) in Bordeaux, Clermont, Marseille, Nancy, Paris, Poitiers, Reims and Strasbourg using prescribed and over the count sales data for asthma (R03) and allergies (R06), from a representative sample of pharmacies (n=12,000), in 2013.

**Results:** In 2013, the 8 cities saw total sales of 9,684,577 R03 and 12,344,102 R06 medications, with sales peaks coinciding with high pollen and air pollution. A spatiotemporal and land cover analysis revealed that relationships between these variables are highly context-dependent. For instance, higher R06 sales clustered in areas with medium pollen (taxon deciles 3 to 4, grass 4 to 5, birch 6 to 7), high pollution (NO_2_ deciles 8 to 10, PM_2.5_ 5 and 8 to 10, PM_10_ 3, 7, 9 to10), high agricultural land, and low forest/urban space. Conversely, some associations did not vary by location, suggesting external influencing factors.

**Conclusions:** Our data raise evidence that pollen and air pollution can act synergistically according to the land cover characteristics. Other investigations are needed to confirm the relationship.

## Introduction

The detrimental impacts of ambient air pollution (NO_2_, PM_2.5_, PM_10_) and airborne pollen (such as birch and grass) on asthma and allergies are well established [1]. Recent evidence also increasingly links the interaction between these factors to these diseases [1].

Indeed, these environmental variables do not act in isolation due to three main mechanisms [3]: 1) Enhanced Allergenicity: Air pollutants can interact directly with airborne pollen grains, causing them to express more allergenic proteins. 2) Deeper Infiltration: While intact pollen grains are often trapped in the upper respiratory tract, they can disintegrate into smaller fragments, attaching to fine particulate matter (PM_2.5_) to penetrate deeper into the lungs. 3) Climate Amplification: Rising temperatures and climate urbanization alter pollination calendars, elongating exposure seasons and inducing plants to produce larger quantities of highly allergenic pollen.

Despite this known synergy, few studies have evaluated the health impacts of the pollution-pollen interaction at a general population level, few population-level studies examine how these dynamics vary across different environments. In a recent systematic review, despite strong evidence from small experimental studies in humans, using common epidemiological study designs only a third of studies identified significant allergen-pollutant interactions [4]. Exposure misclassification, failure to examine subgroups at risk, inadequate statistical power or absence of population-level effects are possible explanations. However, in a population-based time-series study using 20 years of General Practitioner data, co-exposure to high pollen and ozone or PM_2.5_ was associated with elevated allergic rhinitis and asthma risk. [5].

This study addresses this gap by analyzing 2013 medication sales data for allergy and asthma across eight French metropolitan cities representing 13 million people. At the population level, medications sales have been significantly related to both air pollution [6] and pollen [7]. Because sales encompass both prescription and over-the-counter purchases, they serve as a highly exhaustive and statistically powerful proxy for tracking mild-to-severe health exacerbations in relation to pollen, air pollutants, and varying urban, rural, and natural landscapes.

## Materials and Methods

### Data Collection

#### Air Pollution

We obtained outdoor air pollution data, which included daily concentrations (µg/m^3^) of common air pollutants such as NO_2_, PM_10_, and PM_2.5_, from the CHIMERE chemistry-transport model. We were able to pool air pollution data for every 5 km^2^ of space, in a 30 km^2^ area for each of the 8 cities being studied.

#### Pollen

We also obtained daily ambient pollen concentrations (µg/m3) from birch (*Betula*) trees, grass, as well as a “taxon” category including various health-significant pollen, from the RNSA (Le Réseau National de Surveillance Aérobiologique). The spatial resolution acquired for pollen data was for every 30 km^2^ of space, and these values were applied to every 5 km^2^ in a 30 km^2^ area of each city, to render the air pollution and pollen data spatially compatible.

#### Medication sales

We obtained daily data for the sales of medications (per unit sold) used to treat allergies (antihistamines) (R06) and asthma (R03, the category for drugs for obstructive airway diseases, commonly used in epidemiological studies as a proxy for asthma treatment). Medication sales’ data was collected by the Human Data Science Company, IQVIA, from 12,000 pharmacies that are representative of the pharmacies of Metropolitan France, per each 5 km^2^ of space in 8 French metropolitan cities. All data (from CHIMERE, RNSA, and IQVIA) are from the year 2013 and pertain to the following 8 cities in Metropolitan France: Bordeaux, Clermont, Marseille, Nancy, Paris, Poitiers, Reims, and Strasbourg (Figure 1). In addition, we collected land use data from the CORINE Land Cover Database. This data detailed the proportion of each land cover type which consisted of the following categories: urban fabric/artificial surfaces, agriculture, and forest space.

**Figure 1.**
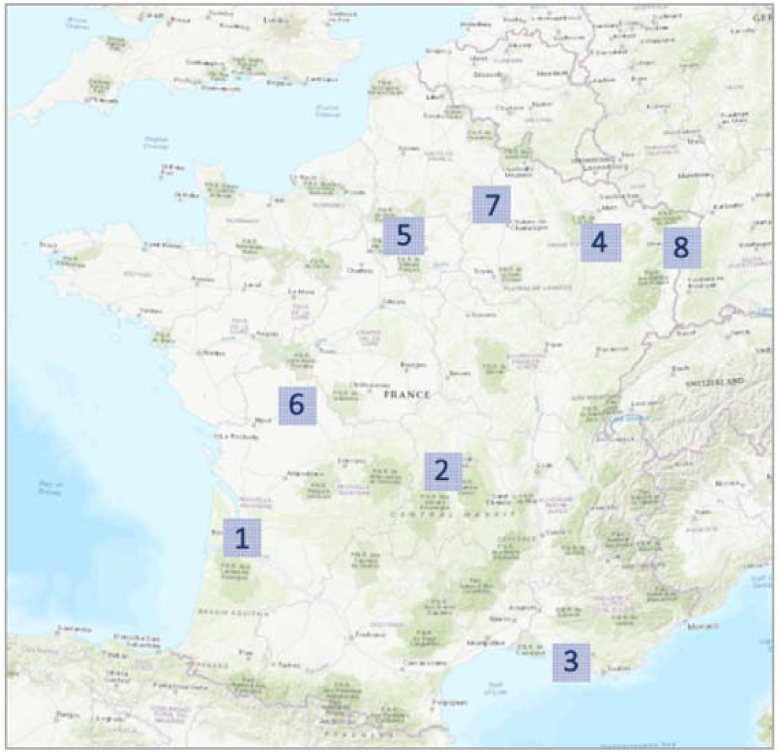
Map of Metropolitan France, depicting the 30 km2 area analyzed in each of the 8 cities: Bordeaux (1), Clermont (2), Marseille (3), Nancy (4), Paris (5), Poitiers (6), Reims (7), Strasbourg (8).

### Data Management

Prior to running analyses on the data, we filtered out observations on dates when pharmacies were closed (this often fell on Sundays and national holidays) because no drugs were sold on these specific dates, although pollution and pollen data was collected. Since the overall aim of this study was to assess the spatio-temporal relationship between all pollen, pollution, and medication sales’ variables, the selection of an optimal spatio-temporal resolution was required before we were able to move forward with advanced statistics. Entries for medication sales in the crude dataset were weighted by population per each 5 km^2^ cell. Then, we standardized all entries for each variable (pollutants, pollen, medication sales) by subtracting the mean from each observation value, and dividing this new value by the standard deviation using the following equation where *X_i_ ‘* equals the standardized feature value, *X_i_* equals the value of the *i^th^* observation, 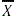 equals the mean value for the variable being assessed, and σ equals the standard deviation for variable being assessed:

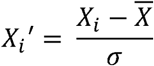

Data were population-weighted and standardized so that ambient air pollution concentrations, airborne pollen concentrations, and medication sales could be spatially comparable across all 5 km^2^ cells and across all 8 cities included in the study. The population-weighted, and standardized, 5 km^2^, daily dataset was aggregated several different ways so we could create the following data frames that vary in spatio-temporal resolutions: 5 km^2^ weekly, 5 km^2^ monthly, 30 km^2^ daily, 30 km^2^ weekly, and 30 km^2^ monthly. To aggregate the population-weighted, standardized, daily dataset into a weekly or monthly dataset, we summarized values for pollutants (NO_2_, PM_10_, and PM_2.5_) using the median of values by week or month, while pollen (birch, grass, and taxon) and medication sales (R03, R06) were summarized using the sum of values by week or month. To aggregate the population-weighted, standardized, 5 km^2^ dataset into a 30 km^2^ dataset, values for all pollutant variables were again summarized using the median of values, values for all pollen variables were summarized using the minimum of all recorded values, and values for all medication sales’ variables were summarized using the sum of all values. We also linked medication sales to pollen and pollution concentrations from 1, 3, and 5 days prior, in order to assess any potential lag effect between the variables, and compared it to the results generated with regular data frame that did not account for a lag effect.

### Statistical Analysis

We executed descriptive statistics on all aggregated data sets (crude, standardized, and lagged: 5 km^2^ daily, 5 km^2^ weekly, 5 km^2^ monthly, 30 km^2^ daily, 30 km^2^ weekly, and 30 km^2^ monthly) to visualize trends, and to decide which resolution best showcased the general trends in the data. Measures of central tendency, such as the mean, median, and mode were calculated, and histograms were generated to visualize the distributions of all variables in each resolution. Time series graphs were generated by plotting pollen, pollutants, and medication sales over time (day/week/month) in both spatial resolutions (5 km^2^ and 30 km^2^) for the year 2013. We also generated scatter plots with respective Pearson correlation coefficients, by plotting the independent variables (pollen and pollutants) against the dependent variables (medication sales) in all spatio-temporal resolutions to visualize any potential existing relationships.

The data used in this ecological study consists of multiple independent variables which may be highly correlated to one another, therefore creating a complex, high-dimensional relationship that may be difficult to efficiently and meaningfully interpret. Principal component analysis (PCA) simplifies these complex, multi-dimensional data into fewer dimensions, while retaining only key and pivotal information that summarizes trends and patterns as well as identifying the most influential variables [8]. The principal components that are retained and summarize general trends in the data tend to be the first few components that explain the largest proportion of the analysis’ variance. We performed a series of PCA’s on all datasets of varying spatio-temporal resolutions (standardized 5 km^2^ daily, 5 km^2^ weekly, 5 km^2^ monthly, 30 km^2^ daily, 30 km^2^ weekly, and 30 km^2^ monthly) and produced correlations of each variable in each dimension, along with respective p-values. We did not additionally manage the data to execute PCA’s, other than weighting the medication sales by population and standardizing all values.

Although PCA aided in dimension reduction and in the generalization of the complex and multi-faceted relationships between all variables, it was not capable of capturing any sort of spatial relationship (e.g. land cover) linked to variables like pollution, pollen, and medication sales. Hierarchical Cluster Analysis (HCA) clusters data into groups based on the similarity or proximity of patterns between values in the data. The overall concept of HCA is such that members of the same cluster are highly associated to each other while being weakly associated to members of other clusters. There are 2 main methods of clustering: agglomerative (bottom-up) and divisive (top-down) clustering [9]. For this study, we utilized an agglomerative clustering, where individual values were clustered to the nearest observation, starting with many clusters until the end-point of the analysis was eventually reached, leaving one remaining cluster (Figure A1).

Prior to HCA execution, we converted the population-weighted and standardized, monthly, 30 km^2^ dataset into a quantitative, ordinal dataset. We categorized all observation values for each of the variables (birch pollen, grass pollen, taxon, NO_2_, PM_10_, PM_2.5_, R03, R06) into decile categories to allow for classification in the HCA. Land cover data was acquired from CORINE Land Cover (CLC) [10]and details the proportion of how land space per each 5 km^2^ cell is utilized in terms of urban/rural space, forests, and agriculture. Land use data variables (urban fabric, agriculture, forests) were each categorized using natural breaks in the distribution to determine how many ordinal groups to generate, so that they could also be subjected to HCA. For urban spaces, we generated 4 ordinal categories (1 = 0 – 0.11, 2 = 0.12 – 0.34, 3 = 0.35 – 0.65, and 4 = 0.66 – 1.5). For agricultural spaces, we generated 3 ordinal categories (1 = 0 – 0.29, 2 = 0.30 – 0.65, and 3 = 0.66 – 1.5). For forest spaces, we generated 3 ordinal categories (1 = 0 – 0.20, 2 = 0.21 – 0.54, and 0.55 = 1.5).

After classification, we merged categorized land use data with the monthly, 5 km^2^, decile data for pollen, pollutants, and medication sales. Since the 4 medication sales’ variables are highly correlated to one another, we conducted 4 separate HCAs, assessing the relationship between pollen, pollution, land use, and the sales of each individual medication type. We determined an optimal number of clusters by using the “elbow method” for each of the 4 HCAs (Figure A2). From this method, we decided on an optimal cluster number of 5 for all 4 HCAs to be executed. After HCAs were completed, dendrogram plots were generated for each HCA, and HCA output statistics were calculated. HCA statistics included the variable composition of each of the 5 clusters, the weight of each variable and its corresponding decile or category in each cluster, p-values, and v-test values.

HCA outputs for R03 and R06 are presented in Tables 3 and 4.

**Table 1.**
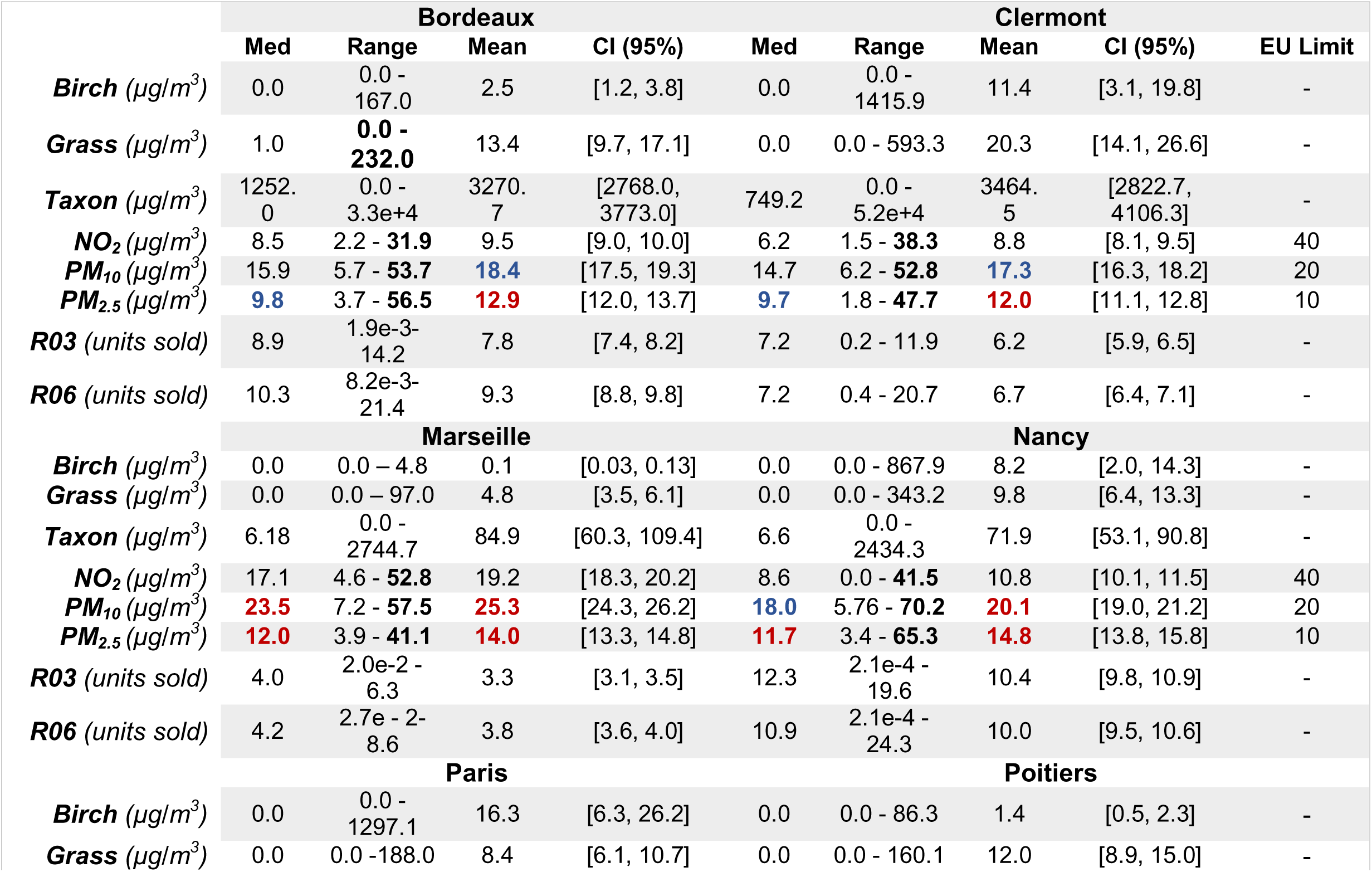
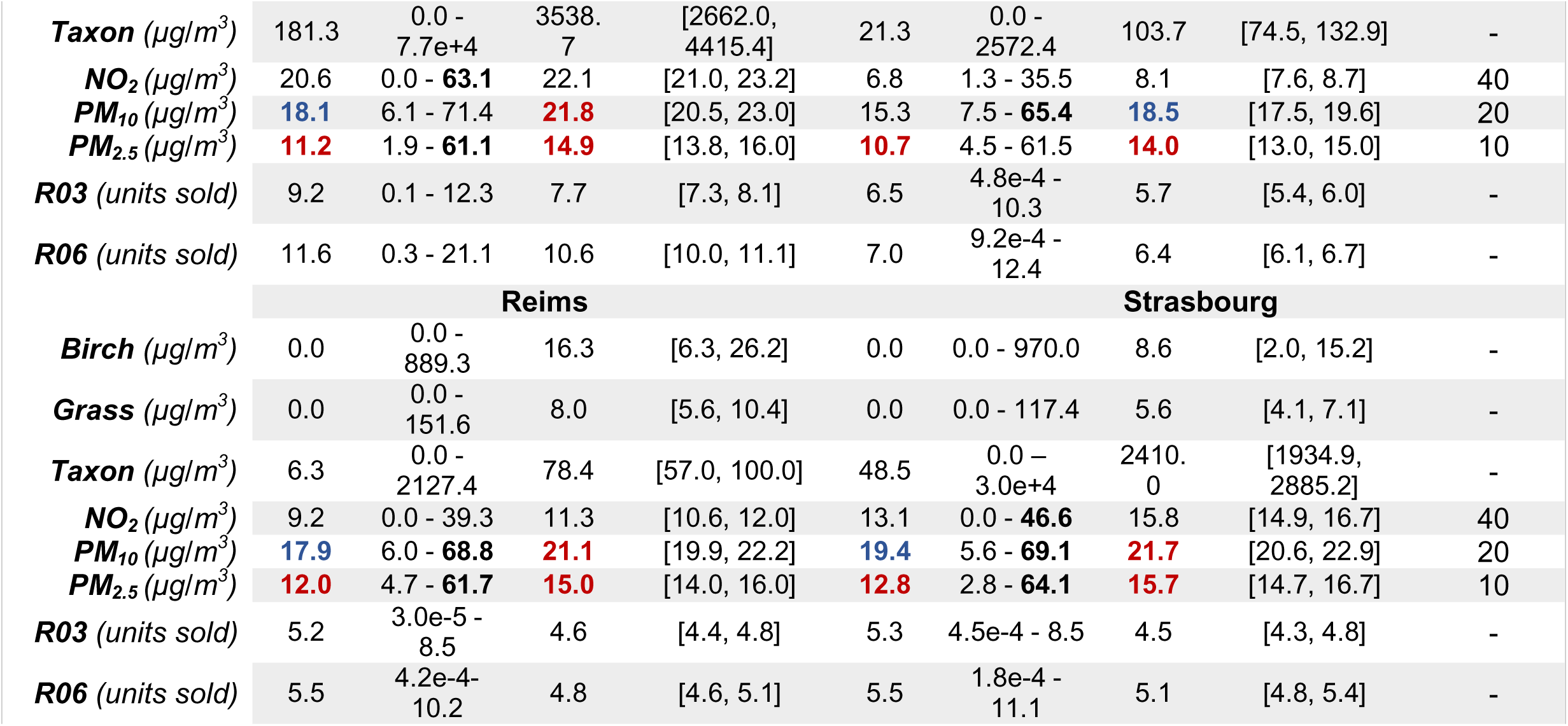
Measures of central tendency calculated for concentration of various pollen and pollutants (µg/m^3^) and sales of medications for asthma, allergies, and sleeping issues/disorders, by city, in France, in 2013. Note: Medication sales are represented as units sold per population of 1000. (R03 = prescription asthma medication, R06 = OTC and prescription allergy medication).

|  | Bordeaux |  |  |  | Clermont |  |  |  | EU Limit |
| --- | --- | --- | --- | --- | --- | --- | --- | --- | --- |
|  | Med | Range | Mean | CI (95%) | Med | Range | Mean | CI (95%) |  |
| **Birch** ($\mu\text{g}/\text{m}^3$) | 0.0 | 0.0 - 167.0 | 2.5 | [1.2, 3.8] | 0.0 | 0.0 - 1415.9 | 11.4 | [3.1, 19.8] | - |
| **Grass** ($\mu\text{g}/\text{m}^3$) | 1.0 | **0.0 - 232.0** | 13.4 | [9.7, 17.1] | 0.0 | 0.0 - 593.3 | 20.3 | [14.1, 26.6] | - |
| **Taxon** ($\mu\text{g}/\text{m}^3$) | 1252.0 | 0.0 - 3.3e+4 | 3270.7 | [2768.0, 3773.0] | 749.2 | 0.0 - 5.2e+4 | 3464.5 | [2822.7, 4106.3] | - |
| **NO<sub>2</sub>** ($\mu\text{g}/\text{m}^3$) | 8.5 | 2.2 - **31.9** | 9.5 | [9.0, 10.0] | 6.2 | 1.5 - **38.3** | 8.8 | [8.1, 9.5] | 40 |
| **PM<sub>10</sub>** ($\mu\text{g}/\text{m}^3$) | 15.9 | 5.7 - **53.7** | **18.4** | [17.5, 19.3] | 14.7 | 6.2 - **52.8** | **17.3** | [16.3, 18.2] | 20 |
| **PM<sub>2.5</sub>** ($\mu\text{g}/\text{m}^3$) | **9.8** | 3.7 - **56.5** | **12.9** | [12.0, 13.7] | **9.7** | 1.8 - **47.7** | **12.0** | [11.1, 12.8] | 10 |
| **R03** (units sold) | 8.9 | 1.9e-3 - 14.2 | 7.8 | [7.4, 8.2] | 7.2 | 0.2 - 11.9 | 6.2 | [5.9, 6.5] | - |
| **R06** (units sold) | 10.3 | 8.2e-3 - 21.4 | 9.3 | [8.8, 9.8] | 7.2 | 0.4 - 20.7 | 6.7 | [6.4, 7.1] | - |
|  | Marseille |  |  |  | Nancy |  |  |  | EU Limit |
|  | Med | Range | Mean | CI (95%) | Med | Range | Mean | CI (95%) |  |
| **Birch** ($\mu\text{g}/\text{m}^3$) | 0.0 | 0.0 - 4.8 | 0.1 | [0.03, 0.13] | 0.0 | 0.0 - 867.9 | 8.2 | [2.0, 14.3] | - |
| **Grass** ($\mu\text{g}/\text{m}^3$) | 0.0 | 0.0 - 97.0 | 4.8 | [3.5, 6.1] | 0.0 | 0.0 - 343.2 | 9.8 | [6.4, 13.3] | - |
| **Taxon** ($\mu\text{g}/\text{m}^3$) | 6.18 | 0.0 - 2744.7 | 84.9 | [60.3, 109.4] | 6.6 | 0.0 - 2434.3 | 71.9 | [53.1, 90.8] | - |
| **NO<sub>2</sub>** ($\mu\text{g}/\text{m}^3$) | 17.1 | 4.6 - **52.8** | 19.2 | [18.3, 20.2] | 8.6 | 0.0 - **41.5** | 10.8 | [10.1, 11.5] | 40 |
| **PM<sub>10</sub>** ($\mu\text{g}/\text{m}^3$) | **23.5** | 7.2 - **57.5** | **25.3** | [24.3, 26.2] | **18.0** | 5.76 - **70.2** | **20.1** | [19.0, 21.2] | 20 |
| **PM<sub>2.5</sub>** ($\mu\text{g}/\text{m}^3$) | **12.0** | 3.9 - **41.1** | **14.0** | [13.3, 14.8] | **11.7** | 3.4 - **65.3** | **14.8** | [13.8, 15.8] | 10 |
| **R03** (units sold) | 4.0 | 2.0e-2 - 6.3 | 3.3 | [3.1, 3.5] | 12.3 | 2.1e-4 - 19.6 | 10.4 | [9.8, 10.9] | - |
| **R06** (units sold) | 4.2 | 2.7e-2 - 8.6 | 3.8 | [3.6, 4.0] | 10.9 | 2.1e-4 - 24.3 | 10.0 | [9.5, 10.6] | - |
|  | Paris |  |  |  | Poitiers |  |  |  | EU Limit |
|  | Med | Range | Mean | CI (95%) | Med | Range | Mean | CI (95%) |  |
| **Birch** ($\mu\text{g}/\text{m}^3$) | 0.0 | 0.0 - 1297.1 | 16.3 | [6.3, 26.2] | 0.0 | 0.0 - 86.3 | 1.4 | [0.5, 2.3] | - |
| **Grass** ($\mu\text{g}/\text{m}^3$) | 0.0 | 0.0 - 188.0 | 8.4 | [6.1, 10.7] | 0.0 | 0.0 - 160.1 | 12.0 | [8.9, 15.0] | - |
| **Taxon** ($\mu\text{g}/\text{m}^3$) | 181.3 | 0.0 - 7.7e+4 | 3538.7 | [2662.0, 4415.4] | 21.3 | 0.0 - 2572.4 | 103.7 | [74.5, 132.9] | - |
| **NO<sub>2</sub>** ($\mu\text{g}/\text{m}^3$) | 20.6 | 0.0 - **63.1** | 22.1 | [21.0, 23.2] | 6.8 | 1.3 - 35.5 | 8.1 | [7.6, 8.7] | 40 |
| **PM<sub>10</sub>** ($\mu\text{g}/\text{m}^3$) | **18.1** | 6.1 - 71.4 | **21.8** | [20.5, 23.0] | 15.3 | 7.5 - **65.4** | **18.5** | [17.5, 19.6] | 20 |
| **PM<sub>2.5</sub>** ($\mu\text{g}/\text{m}^3$) | **11.2** | 1.9 - **61.1** | **14.9** | [13.8, 16.0] | **10.7** | 4.5 - 61.5 | **14.0** | [13.0, 15.0] | 10 |
| **R03** (units sold) | 9.2 | 0.1 - 12.3 | 7.7 | [7.3, 8.1] | 6.5 | 4.8e-4 - 10.3 | 5.7 | [5.4, 6.0] | - |
| **R06** (units sold) | 11.6 | 0.3 - 21.1 | 10.6 | [10.0, 11.1] | 7.0 | 9.2e-4 - 12.4 | 6.4 | [6.1, 6.7] | - |
|  | **Reims** |  |  |  | **Strasbourg** |  |  |  |  |
| **Birch** ($\mu\text{g}/\text{m}^3$) | 0.0 | 0.0 - 889.3 | 16.3 | [6.3, 26.2] | 0.0 | 0.0 - 970.0 | 8.6 | [2.0, 15.2] | - |
| **Grass** ($\mu\text{g}/\text{m}^3$) | 0.0 | 0.0 - 151.6 | 8.0 | [5.6, 10.4] | 0.0 | 0.0 - 117.4 | 5.6 | [4.1, 7.1] | - |
| **Taxon** ($\mu\text{g}/\text{m}^3$) | 6.3 | 0.0 - 2127.4 | 78.4 | [57.0, 100.0] | 48.5 | 0.0 - 3.0e+4 | 2410.0 | [1934.9, 2885.2] | - |
| **NO<sub>2</sub>** ($\mu\text{g}/\text{m}^3$) | 9.2 | 0.0 - 39.3 | 11.3 | [10.6, 12.0] | 13.1 | 0.0 - **46.6** | 15.8 | [14.9, 16.7] | 40 |
| **PM<sub>10</sub>** ($\mu\text{g}/\text{m}^3$) | **17.9** | 6.0 - **68.8** | **21.1** | [19.9, 22.2] | **19.4** | 5.6 - **69.1** | **21.7** | [20.6, 22.9] | 20 |
| **PM<sub>2.5</sub>** ($\mu\text{g}/\text{m}^3$) | **12.0** | 4.7 - **61.7** | **15.0** | [14.0, 16.0] | **12.8** | 2.8 - **64.1** | **15.7** | [14.7, 16.7] | 10 |
| **R03** (units sold) | 5.2 | 3.0e-5 - 8.5 | 4.6 | [4.4, 4.8] | 5.3 | 4.5e-4 - 8.5 | 4.5 | [4.3, 4.8] | - |
| **R06** (units sold) | 5.5 | 4.2e-4 - 10.2 | 4.8 | [4.6, 5.1] | 5.5 | 1.8e-4 - 11.1 | 5.1 | [4.8, 5.4] | - |

**Table 2.**
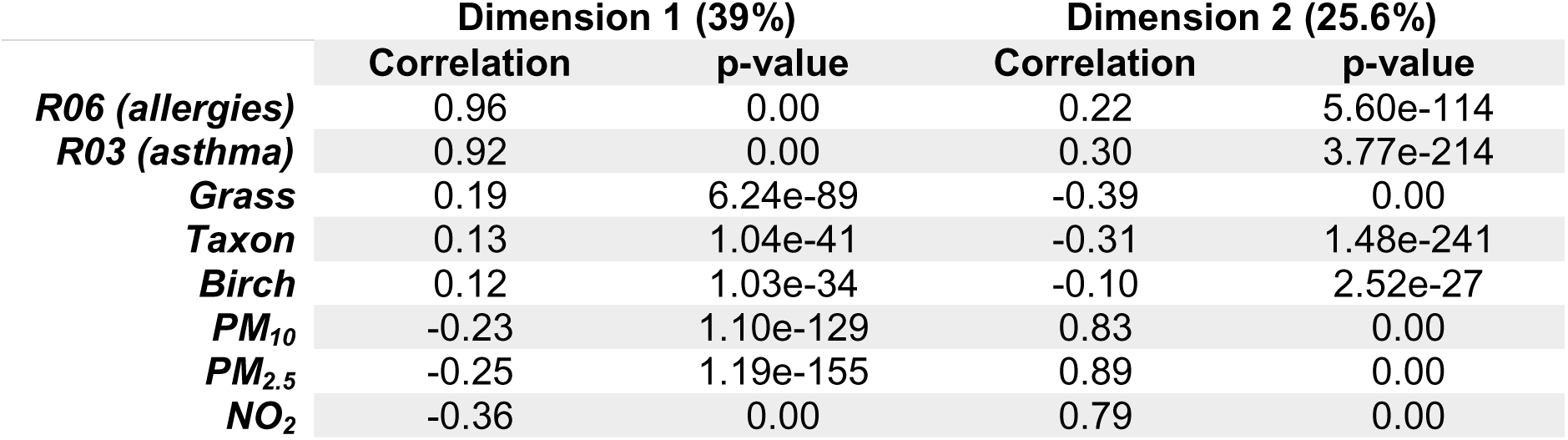
PCA results listing the correlation between each variable being studied and each principal component, in Metropolitan France, in 2013, using a 5 km2, monthly data frame.

**Table 3.** R03 (prescription asthma medication) Hierarchical Cluster Analysis statistics listing variables most associated with each cluster, along with its respective decile or category. “Cla/Mod” represents the percentage of variables with their respective decile/category associated each particular cluster. “Mod/Cla” represents the number of observations in the cluster that are associated with each particular variable and respective decile/category. “Global” represents the percentage of total observations that fall under each specific variable and its respective decile/category.

| Cluster No. | Variable | Decile | Category | Cla/Mod | Mod/Cla | Global | p-value | V-test |
| --- | --- | --- | --- | --- | --- | --- | --- | --- |
| 3 | Taxon | 2 | - | 85.07 | 32.20 | 10.63 | 0.00 | Inf |
| 3 | Grass | 6 | - | 88.43 | 33.03 | 10.49 | 0.00 | Inf |
| 3 | Birch | 10 | - | 79.85 | 27.65 | 9.72 | 0.00 | 36.87 |
| 3 | PM<sub>2.5</sub> | 6 | - | 79.57 | 28.35 | 10.00 | 0.00 | 37.26 |
| 3 | PM<sub>10</sub> | 8 | - | 64.55 | 23.00 | 10.00 | 0.00 | 26.36 |
| 3 | PM<sub>2.5</sub> | 8 | - | 61.29 | 21.83 | 10.00 | 0.00 | 24.04 |
| 3 | Agriculture | - | 2 | 34.28 | 35.96 | 29.45 | 0.00 | 9.13 |
| 3 | Forest | - | 2 | 33.54 | 37.45 | 31.35 | 0.00 | 8.43 |
| 3 | Urban | - | 1 | 28.96 | 66.63 | 64.61 | 0.01 | 2.74 |
| 3 | R03 | 8 | - | 31.16 | 11.10 | 10.00 | 0.02 | 2.34 |
| 3 | R03 | 10 | - | 31.06 | 11.07 | 10.00 | 0.02 | 2.27 |
| 3 | R03 | 4 | - | 30.88 | 11.00 | 10.00 | 0.03 | 2.13 |
| 5 | Taxon | 3 | - | 78.46 | 61.96 | 10.66 | 0.00 | Inf |
| 5 | Grass | 5 | - | 85.80 | 50.55 | 7.95 | 0.00 | Inf |
| 5 | Grass | 4 | - | 58.09 | 47.44 | 11.02 | 0.00 | Inf |
| 5 | NO<sub>2</sub> | 9 | - | 38.10 | 28.22 | 9.99 | 0.00 | 21.64 |
| 5 | PM<sub>2.5</sub> | 10 | - | 36.19 | 26.83 | 10.00 | 0.00 | 20.12 |
| 5 | PM<sub>10</sub> | 10 | - | 33.21 | 24.62 | 10.00 | 0.00 | 17.67 |
| 5 | Forest | - | 1 | 17.24 | 68.95 | 53.98 | 0.00 | 12.45 |
| 5 | Urban | - | 1 | 16.39 | 78.49 | 64.61 | 0.00 | 12.27 |
| 5 | Agriculture | - | 2 | 11.44 | 24.97 | 29.45 | 0.00 | -4.07 |
| 5 | R03 | 10 | - | 17.82 | 13.21 | 10.00 | 0.00 | 4.22 |
| 5 | R03 | 9 | - | 15.97 | 11.83 | 9.99 | 0.01 | 2.45 |
| 5 | R03 | 1 | - | 11.57 | 8.58 | 10.00 | 0.05 | -1.97 |

**Table 4.** R06 (prescription and OTC allergy medication) Hierarchical Cluster Analysis statistics listing variables most associated with each cluster, along with its respective decile or category. “Cla/Mod” represents the percentage of variables with their respective decile/category associated each particular cluster. “Mod/Cla” represents the number of observations in the cluster that are associated with each particular variable and respective decile/category. “Global” represents the percentage of total observations that fall under each specific variable and its respective decile/category.

| Cluster No. | Variable | Decile | Category | Cla/Mod | Mod/Cla | Global | p-value | V-Test |
| --- | --- | --- | --- | --- | --- | --- | --- | --- |
| 3 | Taxon | 2 | - | 86.30 | 31.67 | 10.63 | 0.00 | Inf |
| 3 | Grass | 6 | - | 89.06 | 32.25 | 10.49 | 0.00 | Inf |
| 3 | Birch | 10 | - | 80.23 | 26.93 | 9.72 | 0.00 | 36.40 |
| 3 | PM<sub>2.5</sub> | 6 | - | 79.38 | 27.42 | 10.00 | 0.00 | 36.35 |
| 3 | PM<sub>10</sub> | 8 | - | 65.30 | 22.55 | 10.00 | 0.00 | 26.14 |
| 3 | PM<sub>2.5</sub> | 8 | - | 61.94 | 21.39 | 10.00 | 0.00 | 23.76 |
| 3 | Agriculture | - | 2 | 34.54 | 35.12 | 29.45 | 0.00 | 8.14 |
| 3 | Forest | - | 2 | 33.93 | 36.73 | 31.35 | 0.00 | 7.59 |
| 3 | Forest | - | 1 | 27.75 | 51.71 | 53.98 | 0.00 | -3.01 |
| 3 | R06 | 8 | - | 34.42 | 11.89 | 10.00 | 0.00 | 4.09 |
| 3 | R06 | 9 | - | 32.68 | 11.28 | 9.99 | 0.01 | 2.80 |
| 3 | R06 | 10 | - | 32.18 | 11.11 | 10.00 | 0.02 | 2.43 |
| 4 | Taxon | 3 | - | 78.46 | 62.01 | 10.66 | 0.00 | Inf |
| 4 | Grass | 5 | - | 85.92 | 50.66 | 7.95 | 0.00 | Inf |
| 4 | Grass | 4 | - | 58.26 | 47.61 | 11.02 | 0.00 | Inf |
| 4 | Birch | 7 | - | 68.99 | 56.82 | 11.10 | 0.00 | Inf |
| 4 | Birch | 6 | - | 74.01 | 34.88 | 6.35 | 0.00 | 38.07 |
| 4 | NO<sub>2</sub> | 9 | - | 38.10 | 28.24 | 9.99 | 0.00 | 21.66 |
| 4 | PM<sub>2.5</sub> | 10 | - | 36.19 | 26.85 | 10.00 | 0.00 | 20.13 |
| 4 | PM<sub>10</sub> | 10 | - | 33.21 | 24.64 | 10.00 | 0.00 | 17.68 |
| 4 | Agriculture | - | 3 | 19.98 | 66.37 | 44.79 | 0.00 | 17.74 |
| 4 | Forest | - | 1 | 17.20 | 68.86 | 53.98 | 0.00 | 12.37 |
| 4 | Urban | - | 1 | 16.32 | 78.20 | 64.61 | 0.00 | 12.00 |
| 4 | R06 | 8 | - | 10.63 | 7.89 | 10.00 | 0.00 | -2.95 |

We then utilized these output HCA statistics to visualize how these clusters were distributed over space and time using ArcGIS software (ArcGIS Pro 2.3.3), creating 3-dimensional spatio-temporal cubes for each city being analyzed (Figure 2). We assessed an area of 30 km^2^ surrounding the city center in each of the 8 cities. We then overlaid these spatio-temporal cubes on a map of Metropolitan France, with a base map showing the distribution of different types of land cover through 5 colors, again using the CORINE Land Cover data, with each spatio-temporal cube corresponding to its respective city. Horizontal dimensions X and Y of the cube use individual cells to describe space per 5 km^2^, with the entire cube representing a 30 km^2^ region of the city. The final dimension of the cube describes time on a vertical Z axis, on a monthly basis for one entire year, where the single 5 km^2^ cubes located on the lowest vertical level represent the month of January, and the single 5 km^2^ cubes on the highest vertical level represent the month of December. The color of each single 5 km^2^ cube corresponds to the cluster number it belongs to, in order to visualize any spatiotemporal trends between pollen, pollution, land use, and medication sales per 5 km^2^ in all 8 French Metropolitan cities, per month, in the year 2013. Figure 3 is a visual example explaining the concept of spatiotemporal cubes taking into account air pollution, pollen, land cover and medications sales as a result of clustering analysis.

**Figure 2.**
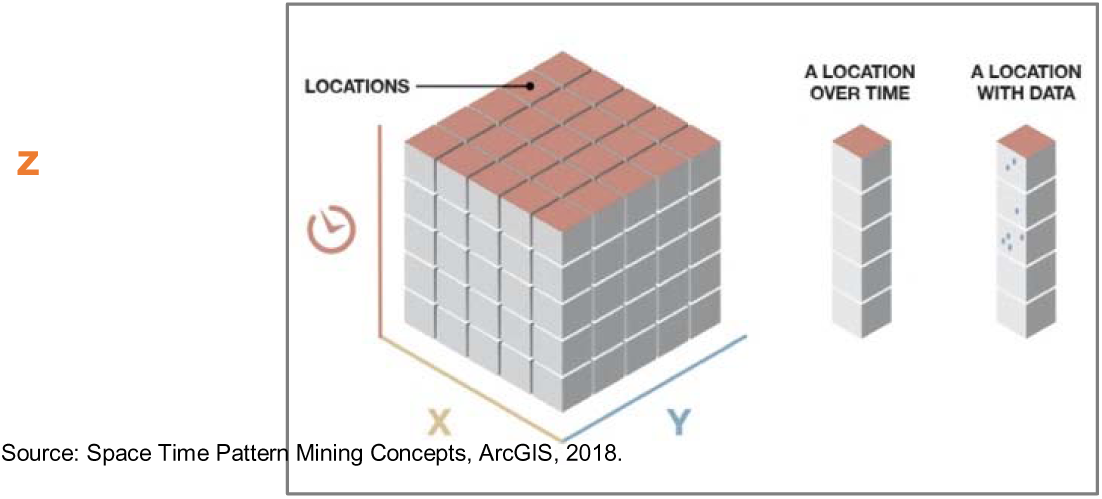
Visual representation of the concept behind a spatio-temporal cube where horizontal axes x and y represent space per 5 km^2^, and vertical axis z represents time per month.

**Figure 3.**
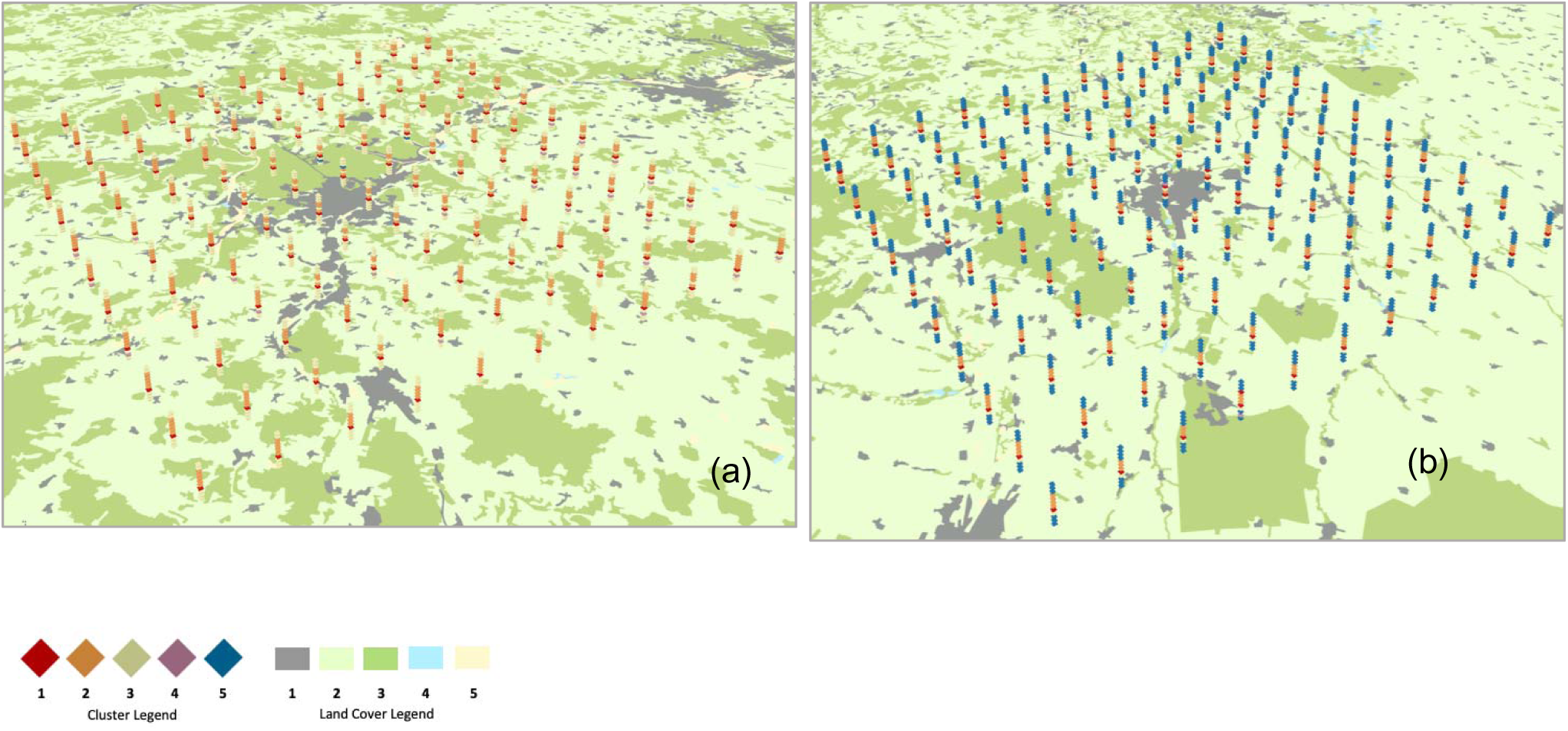
30 km^2^ spatio-temporal cubes for R03 (asthma medication) HCA. **12a**). R03 HCA in a 30 km^2^ region of Nancy, in 2013. **12b).** R03 HCA in a 30 km^2^ region of Reims, in 2013. (Land Cover Legend: 1 = Artificial surfaces, including urban fabric. 2 = Agricultural areas, including crops and pastures. 3 = Forest and semi-natural areas. 4 = Wetlands. 5 = Bodies of Water).

**Figure 12.**
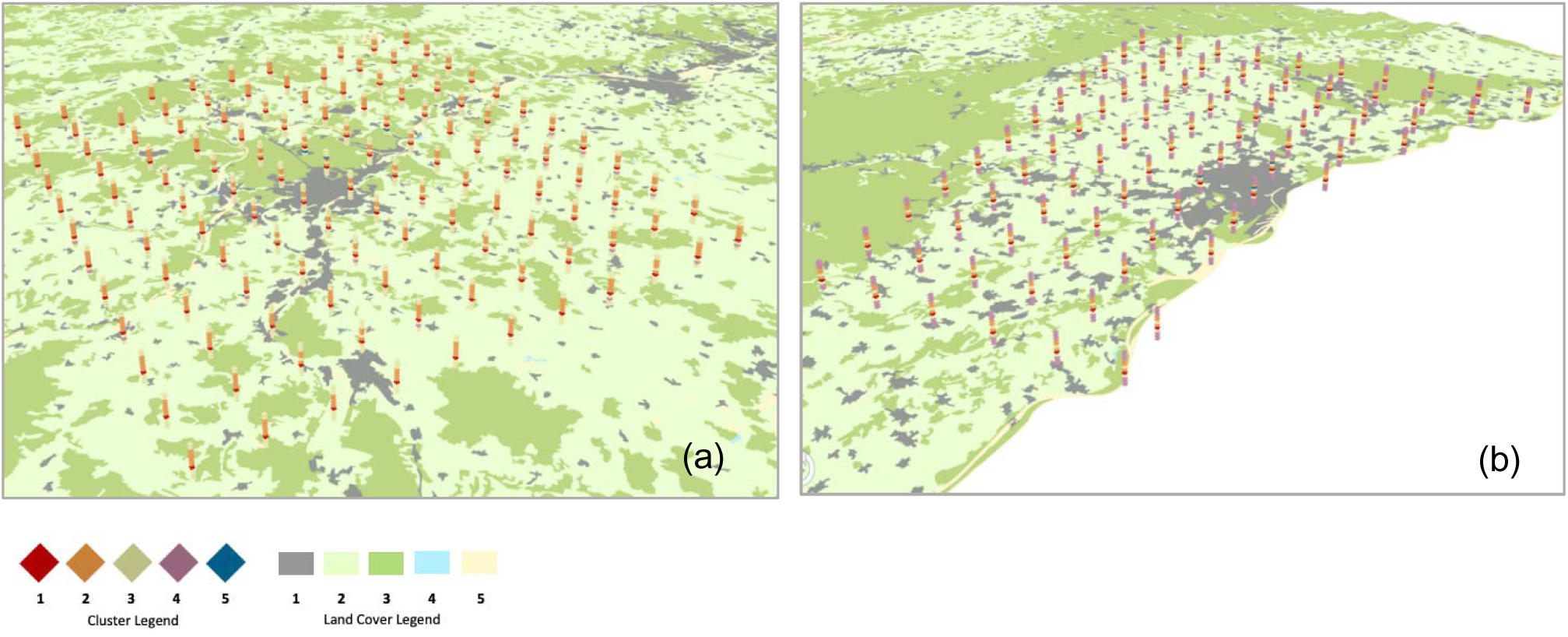
30 km^2^ spatio-temporal cubes for R06 HCA. **12a).** R06 HCA in a 30 km^2^ region of Nancy, in 2013. **12b).** R06 HCA in a 30 km^2^ region of Strasbourg, in 2013. (Land Cover Legend: 1 = Artificial surfaces, including urban fabric. 2 = Agricultural areas, including crops and pastures. 3 = Forest and semi-natural areas. 4 = Wetlands. 5 = Bodies of Water).

## Results

### Distribution of Pollen, Air Pollutants, and Land Cover Characteristics (Measures of Central Tendency)

Overall, there was a total of 9,684,577 R03, and 12,344,102 R06 medications sold in Metropolitan France, in 2013.

The spatio-temporal resolution that best showcased trends during descriptive statistics was that of a 30 km^2^ monthly spatio-temporal resolution. Although medication sales were population-weighted, and all variables were standardized, we observed a large variability across all 8 cities. Therefore, it was not appropriate to generalize the measures of central tendency into one value for each variable, for France as a whole. The median, range, mean, and 95% confidence intervals (CI) for pollen, pollutants, and medication sales, as well as WHO guidelines for pollution limits in the European Union (EU) are reported in Table 1, by variable, for each of the 8 French cities. The values shown in Table 1, for variables such as birch, grass, taxon, NO_2_, PM_10_, and PM_2.5_ are crude concentrations, comparable with EU limits for pollutants, and summarized from a daily, 30 km^2^ data frame. Medication sales have been population-weighted and are reported as units sold per population of 1000. Table 1 shows that all cities have high levels of PM pollution. All 8 cities had an annual mean of PM_2.5_ concentration higher than the 10 µg/m^3^ concentration EU limit suggested by the WHO in 2013. In 2013, Marseille, Nancy, Paris, Reims, and Strasbourg all had an average annual mean of a PM_10_ concentration higher than the 20 µg/m^3^ WHO recommendation, while Bordeaux, Clermont, and Poitiers had PM_10_ concentrations considerably close to this limit (18.4 µg/m^3^, 17.3 4 µg/m^3^, and 18.5 4 µg/m^3^, respectively). No cities had mean annual NO_2_ concentrations higher, or close to the 40 µg/m^3^ WHO recommendation. However, the ranges for NO_2_ concentrations throughout the year, indicate that cities such as Marseille, Nancy, Paris, Poitiers, Reims, and Strasbourg experience concentrations that well surpass this limit during certain days of the year. We also observed large ranges for concentrations of pollen throughout the year. In the most extreme case, Paris had a taxon pollen concentration range of 0.0 to 7.7e+04 µg/m^3^, indicating that pollen could be highly present during certain days of the year, and entirely absent during other days of the year.

We generated histograms for each variable, using the population-weighted, standardized, 30 km^2^, monthly data frame and observed that the distribution of relative concentration for pollen and pollutants, as well as the distribution of relative sales of medications followed the same general distribution according to the class of variable they belonged to (pollen, pollutant, or medication sales). The distribution of the relative concentration of taxon pollen (Figure 1a) is positively skewed, and all other pollen have a similar distribution. The distribution of the relative concentration of PM_2.5_ is also positively skewed.

All other pollutants follow a similar distribution patter for concentration. The distribution of the relativsales of R06 is slightly negatively skewed, but follows more of a normal distribution when compared to the distributions of pollen and pollutant concentrations. The remaining medications (R03) follow a similar relative sales’ distribution.

The distributions regarding the proportions of each type of land cover were also examined and we found that low proportions were the most frequent in urban spaces, resulting in a positive distribution skew. For agricultural spaces, high proportions were the most frequent, and the distribution was therefore negatively skewed. Forest space proportion distribution was positively skewed due to a higher frequency of lower proportions. We did not observe any significant differences between the lagged and unlagged data sets, so only regular, unlagged data and results will be discussed moving forward.

### Time Evolution

To evaluate the daily and seasonal trends of aeroallergen emissions, ambient pollutant concentrations, and corresponding pharmaceutical sales, temporal analyses were conducted using standardized daily data at a 30 km resolution. This resolution optimized the capture of distinct seasonal successions and annual baselines across the studied metropolitan areas, using Paris as a representative urban template. The temporal tracking of airborne pollen revealed strict, non-overlapping seasonal successions characterized by acute spikes and long periods of total latency (data not shown): Birch Pollen exhibited the most rigid seasonality, peaking sharply between late April and early May, before immediately dropping to a absolute baseline of zero for the remainder of the year. Grass Pollen demonstrated a distinct summer seasonality, rising in June and progressively declining to zero by August. Taxon Pollen showed a more protracted footprint in the warmer months, climbing in July, maximizing during August and September, and returning to a zero baseline before October. Cumulatively, these data confirm that allergenic exposure is absent during late autumn and winter, concentrated exclusively in high-amplitude spikes during spring and summer.

In stark contrast to the binary presence of pollen, ambient air pollutants (NO_2_ and PM_2.5_) maintained a persistent, chronic baseline throughout the year (data not shown). Both pollutants demonstrated a strong winter-dominant profile, with relative concentrations maximizing from November through March. This pattern aligns with increased anthropogenic emissions, such as residential heating, during colder periods. However, fine particulate matter (PM_2.5_) sustained sharp, episodic concentration spikes into April. While pollution levels declined during the spring, summer, and autumn, they never leveled off to a flat baseline in the manner observed with aeroallergens.

### Converting observations of possibly correlated variables (Principal Component Analysis)

Since the multiple independent variables being studied (pollen and air pollutants) could be highly correlated with each other, we executed several PCA’s using varying spatio-temporal resolutions to observe the relationships with only the necessary dimensions. Tables 2 lists the correlation each variable has with each principal component in the analyses. This allowed to better perform the rest of analysis.

### Spatiotemporal Characterization via Hierarchical Cluster Analysis

To evaluate the potential synergistic effects of co-exposure to ambient air pollution and airborne pollen across varying landscapes, Hierarchical Cluster Analysis (HCA) was executed separately for prescribed asthma medications (R03) and allergy medications (R06) at a 5 km monthly resolution. A five-cluster solution was applied to each model. To isolate the most epidemiologically relevant signals, the analysis was restricted to clusters demonstrating concurrent medium-to-high exposure deciles for both pollen and pollution alongside significant associations with medication sales; clusters capturing opposing environmental trends or lacking clear sales associations were omitted.

<u>Prescribed Asthma Medications</u> (R03) (Table 3)

Two distinct co-exposure clusters emerged as statistically relevant for asthma medication sales, revealing highly individualized geographical and seasonal variations across the eight metropolitan areas:

- Cluster 3 (Moderate Co-Exposure of pollen and air pollution, Low Urbanization): Characterized by medium-to-high airborne pollen, medium-to-high ambient air pollution (PM_2.5_, PM_10_, and NO_2_), mild agricultural and forest cover, and low urban space. This profile was associated with low-to-medium R03 sales.
- Cluster 5 (High Co-Exposure of pollen and air pollution, High Agriculture): Defined by medium-to-high pollen loads, high pollution concentrations, high agricultural land cover, and low urban density. This profile exhibited a polarizing relationship with health outcomes, showing strong associations with both extremely high and low R03 sales deciles.

The spatial and temporal manifestation of these profiles split the studied cities into distinct cohorts. In Bordeaux, Marseille, and Poitiers, Cluster 3 was uniformly distributed across space but strictly bounded in time, capturing nearly all spatial cells during March and April, while Cluster 5 was entirely absent. Conversely, in Paris, Cluster 3 predominated during the middle of the year, spanning February to July in the peripheral zones but contracting to an un-uniform footprint from April to July in the urban core.

A separate spatiotemporal pattern emerged in Clermont and Nancy, where Cluster 3 persisted during the colder months (October through February), shifting closer toward the urban center in Nancy. Cluster 5 appeared rarely in these locations, confined to the northeast and central sectors of Clermont during February and September, and restricted to January in Nancy. In contrast, Reims and Strasbourg demonstrated a profound temporal dependency rather than spatial variation; Cluster 5 uniformly covered both cities from September through April, while Cluster 3 uniformly emerged to represent the month of April.

Figure 2 depicts the situation for allergy medication sales in Nancy and Reims respectively. Cluster 5 (blue) is rare in the former but frequent in some months of the latter.

<u>Prescribed and Over-the-Counter Allergy Medications</u> (R06)

The allergy medication HCA similarly identified two profiles marked by concurrent environmental stressors, both strongly associated with elevated medication demand:

- Cluster 3 (Moderate Balanced Co-Exposure): Characterized by moderate levels of both airborne pollen and ambient pollutants, alongside a mild presence of agricultural and forest landscapes. This cluster was strongly linked to high-level R06 sales.
- Cluster 4 (High Pollution Co-Exposure): Defined by moderate pollen levels paired with high air pollution deciles, high agricultural land use, and low forest and urban space. This profile was uniquely tied to exceptionally high R06 sales.

Geographically, the distribution of these allergy-related profiles was highly region-specific. In Bordeaux, Paris, and Poitiers, Cluster 3 displayed complete spatial uniformity, consistently defining the months of March, April, and July across all peripheral and central grids, whereas Cluster 4 did not appear. Marseille exhibited a similarly uniform but brief footprint, where Cluster 3 appeared across all grids but was strictly limited to a single month during the spring or summer season.

A shifting pattern was documented in Clermont and Nancy, where Cluster 3 uniformly spanned from October through February, as well as May and June. Cluster 4 occurred dynamically near the urban centers of these two cities, restricted exclusively to the month of January. Finally, in Reims and Strasbourg, Cluster 4 predominated continuously and uniformly across all spatial cells from August through March, signifying a prolonged, high-pollution environmental matrix that strongly drives population-level allergy exacerbations throughout the autumn and winter months.

Figure 3 depicts the situation for allergy medication sales in Nancy and Reims respectively. This time, cluster 5 (blue) is rare in both situations.

## Discussion

Using total medication sales as a highly sensitive proxy for population-level disease activity, this ecological analysis reveals how varying landscapes alter the real-world impact of environmental stressors like air pollution and pollen on asthma and allergy medications sales.

For the first time, our study assessed the combined impact of the complex interrelationships among ambient air pollution, airborne pollen, and land cover characteristics on national asthma and allergy medication sales. Our findings underscore the importance of considering the spatiotemporal dimension of environmental exposures, as both air pollution and pollen concentrations exhibit marked spatial and seasonal variability. These factors not only co-vary over time and across locations but also interact with land cover and meteorological conditions, shaping population exposure and, ultimately, the burden of allergic and respiratory diseases.

Highly distinct temporal and spatial profiles for ambient air pollutants (NO_2_, PM_2.5_, PM_10_) versus aeroallergens (birch, grass, and taxon pollen) across eight metropolitan areas in France, representing 13 million inhabitants, were observed. Ambient air pollution maintained a chronic, year-round baseline presence that peaked during winter months. While annual mean NO_2_ concentrations remained below the 2005 World Health Organization (WHO) threshold (40.0 g/m^3^), short-term seasonal spikes frequently exceeded this limit. Alarmingly, all eight cities exhibited mean and median fine particulate matter (PM_2.5_) concentrations that consistently violated the WHO public health guideline (10 g/m^3^), indicating chronic population exposure to hazardous particulate levels.

Conversely, pollen emissions were characterized by acute, transient, and highly localized spikes during spring and summer. Birch pollen displayed the most extreme seasonality, peaking sharply around May and remaining entirely absent for the rest of the year. The magnitude of these exposures varied vastly by geography; for instance, maximum annual birch pollen concentrations reached 167.0 g/m^3^ in Bordeaux compared to 1,415.9 g/m^3^ in Clermont. Because air pollution and pollen load exhibit opposing seasonal maxima (winter vs. spring/summer), evaluating their synergistic public health impact requires sophisticated spatiotemporal modeling capable of untangling asynchronous windows of exposure.

Hierarchical Cluster Analysis (HCA) and spatial mapping confirmed that regional medication sales closely mirrored environmental fluctuations. Allergy medication (R06) purchases exhibited bimodal seasonal peaks that coincided precisely with May birch and July grass pollen surges. Meanwhile, prescribed asthma medications rose significantly during the winter, establishing a robust positive correlation with elevated NO_2_ and PM concentrations. Notably, a uniform, drastic decline in all medication sales was observed across all eight cities during August. This anomaly reflects a socio-behavioral confounder—the traditional French summer vacation period, during which synchronous drops in active pharmacy customers and prescribing physicians occur—rather than a true reduction in environmental risk.

When modeling space, time, and land cover simultaneously, the relationships proved remarkably non-linear and context-dependent. The baseline hypothesis—that the intersection of high pollution and high pollen loads would uniformly exacerbate health outcomes in highly urbanized spaces—held true only in specific geographical clusters and during restricted periods of the year. For example, a prominent cluster characterized by high agricultural land use alongside low forest and urban space demonstrated an increased vulnerability to R06 sales under conditions of high pollution and moderate pollen deciles.

In other instances, the relationship was independent of urban spatial configurations. This spatial divergence strongly underscores the influence of broader, unmeasured factors within the human exposome – such as localized microclimates, terrain, and overlapping winter influenza epidemics – which complicate the causal pathways between ambient exposures and health outcomes.

### Strengths and Limitations

One of the major advantages of the nature of this ecological study is that we utilized data that already existed and that is continuously being collected (pollen and pollution concentrations, and medication sales from pharmacies). Therefore, the study can be repeated in future years or for an extended period of time, rather than just 1 year. This study also considered that many previous studies have attempted to assess the health impacts of pollen and pollution with regards to asthma and allergies by observing hospital admissions and emergency room visits, which only factors extreme cases of asthma attacks. Using allergy, asthma, and sleep medication sales as a proxy indicator is more statistically powerful, as it increases the sample size, and also captures less-severe cases of asthma and allergic disease. This study included both prescription and over the counter medication, which is more exhaustive and captures even more cases.

There were some limitations to the study that might account for the lack of clear association, and that should be considered in future studies. Perhaps the year selected for this study, 2013, was not the best representation of the situation in Metropolitan France. In the future, this study could be conducted using data from other years and compare findings across different years to see if the same results are observed. We also often encountered the issue of potential exposure misclassification as the resolution of pollen assessment was too broad. Additionally, pollen concentrations were very low or absent for most of the year, making it difficult to analyze the interaction between pollen and air pollutants. Perhaps future studies could select a location where both pollen and pollution concentrations are assessed at higher resolution so to increase statistical power. Another limitation depends on the fact that we used an ecological approach, as considered as the most pertinent based on available data. Therefore it is difficult to disentangle the health impacts of each individual pollutant and it is difficult to conclude which pollutant poses the greatest health risk, as it is mostly a collective effect. In fact, some studies argue that although NO_2_ is a common indicator used to quantify air pollution, its health impacts might be confounded by its strong correlation with other air pollutants. NO_2_ is a weak oxidant, causing some debate in the scientific community regarding whether or not it is directly associated with asthma morbidity and mortality [15].

Another limitation in this study is that the data we acquired for medication purchases does not include personal data about the patient allowing adjustment. No data was available regarding sex or age of the patient; therefore, we could not stratify our analyses by certain demographics to identify vulnerable populations. Also, we could not adjust for smoking that is related to asthma and sleeping disorders. Data regarding how many boxes of medications were purchased by the same person was also not available, which might have proven to be useful if patients were buying both asthma and allergy medications at the same time during periods where pollen and pollution concentrations were elevated. Furthermore, this study was unable to account for the individuals who purchased medications in bulk to last them for a longer period, and we could therefore not determine when these medications were being taken, the dose being taken, and if this was related to pollen or pollution concentrations.

This study did not factor or consider meteorological conditions or flu season data, which could be of relevance in future studies. Finally, this study heavily relied on France’s centralized healthcare system where sales of medications are automatically recorded and logged onto an electronic database. This study is not reproducible in countries where this type of sales’ logging system is not already in place, which might leave out important data from countries that are most vulnerable to the effects of air pollution and pollen on public health.

### Conclusion

These findings carry vital public health urgency in the context of anthropogenic climate change. Global warming is actively destabilizing atmospheric chemistry and modifying plant biology – resulting in elongated pollination calendars, higher volumes of allergen release, and amplified allergenicity driven by direct chemical interactions with ambient pollutants. Because the real-world impact of these combined environmental stressors is mediated by local land cover and seasonal shifts, public health interventions and climate adaptation frameworks cannot rely on broad national mandates. Instead, mitigation policies must be dynamically tailored to the unique spatiotemporal and environmental landscapes of individual urban and rural regions.

## Data Availability

Contact the corresponding author

## Acknowledgement

IAM thank Karen Hernandez Dominguez and Julie Prud’homme who worked on this project in the EPAR Dept, IPLESP INSERM AND SORBONNE UNIVERSITY that does not exist anymore. The project was implemented in the frame of The POLLAR: Impact of air POLLution on Asthma and Rhinitis; a European Institute of Innovation and Technology Health (EIT Health) project.

## List of Acronyms and Abbreviations

CH_4_: Methane
CI: Confidence Interval
CO_2_: Carbon Dioxide
CLC: CORINE Land Cover
DALY: Disability-Adjusted Life Years
EEA: European Environment Agency
ER: Emergency Room
EU: European Union
HCA: Hierarchical Cluster Analysis
IgE: Immunoglobulin E
IQVIA: an American-based company that provides services in health information and clinical research
NASA: the National Aeronautics and Space Administration
NO_2_: Nitrogen Dioxide
N_2_O: Nitrous Oxide
O_3_: Ozone
OTC: Over the Counter
PCA: Principal Component Analysis
PM: Particulate Matter
POLLAR: Impact of POLLution on Asthma and Rhinitis
R: Correlation coefficient (Pearson)
RNSA: Le Réseau National de Surveillance Aérobiologique (National Aerobiological Surveillance Network)
R03: Class of prescribed medications for obstructive air way diseases
R06: Class of prescribed and over the counter medications for allergies (anti-Histamines for systemic use)
SO_2_: Sulfur Dioxide
WHO: the World Health Organization

## Notes

### Competing Interest Statement

The authors have declared no competing interest.

